# EpidBot: A Natural Language Platform for Generalized Epidemic Intelligence

**DOI:** 10.64898/2026.06.18.26355985

**Authors:** Flávio Codeço Coelho, Joyce Figueiró Braga, Beatriz Laiate

## Abstract

Public health professionals have access to more data than ever before. Yet answering a relatively simple epidemiological question often requires navigating multiple databases, formats, software tools, and reporting systems. As a result, valuable data often remain locked behind technical barriers, making it harder for public health professionals to turn information into decisions.

We developed EpidBot to simplify this process. EpidBot is a platform that allows users to retrieve, analyze, visualize, model, and report epidemiological data through natural language interaction. By connecting multiple public health data sources within a single environment, the platform enables users to conduct analyses that would traditionally require several independent tools and specialized technical skills.

Rather than functioning solely as a search interface, EpidBot supports complete analytical workflows. Users can explore surveillance data, compare trends across locations and time periods, generate maps and visualizations, construct epidemiological models, and produce structured technical reports while maintaining full visibility of data sources and analytical procedures.

To show what this looks like in practice, we present representative use cases, including the automatic generation of a mathematical model for Ebola virus disease in the Democratic Republic of the Congo. From a single user request, EpidBot assembled evidence from published sources, generated and calibrated a compartmental transmission model, identified key transmission drivers, evaluated intervention scenarios, and produced a technical report with quantitative findings and policy-relevant recommendations.

EpidBot shows how natural language interaction can reduce the technical barriers that often separate public health professionals from the analyses they need to perform. By bringing data access, analysis, modeling, visualization, and reporting into a single environment, the platform helps transform information into evidence while preserving transparency and reproducibility.

**Author summary:** Public health professionals rarely struggle with a lack of data. More often, they struggle with the effort required to find the right data, combine information from different sources, perform analyses, create visualizations, and communicate results. Even relatively simple questions can require moving between multiple databases, software tools, and reporting systems.

We created EpidBot to simplify this process. EpidBot allows users to interact with epidemiological data using natural language while preserving the analytical rigor required for public health practice. Instead of serving only as a search tool, the platform supports complete workflows that may include data retrieval, visualization, statistical analysis, mathematical modeling, and report generation.

In this article, we show how these workflows can be applied in practice through a series of examples, including the automatic construction of an Ebola transmission model for the Democratic Republic of the Congo. Starting from a single user request, EpidBot gathered evidence from published sources, identified key transmission drivers, evaluated intervention scenarios, and generated a technical report with quantitative results and public health recommendations.

EpidBot was designed for epidemiologists, surveillance teams, researchers, and public health professionals who need answers, not just data. By reducing technical barriers and bringing multiple analytical tasks into a single environment, the platform helps transform information into evidence that can support public health decisions.

## Introduction

Society is undeniably in the era of abundant health data. Disease notifications, mortality records, hospital admissions, vaccination data, climate indicators, genomic surveillance, and international health statistics are increasingly available through public repositories and digital platforms [1]. Paradoxically, answering a relatively simple epidemiological question, such as predicting the next outbreak of a specific disease, remains a surprisingly difficult achievement [2].

The challenge is rarely the absence of data. More often, it is the effort required to find relevant information, combine data from different sources, perform analyses, generate visualizations, and communicate results. In practice, epidemiologists frequently move among multiple databases, dashboards, spreadsheets, statistical software packages, and reporting tools before producing evidence to supports a decision [3].

This fragmentation creates a gap between data availability and public health action. While surveillance systems continue to generate increasing amounts of information [4], extracting useful insights often requires a combination of epidemiological expertise and technical skills in database querying, programming, data management, statistical analysis, modeling, and visualization. Acquiring and maintaining these skills can be challenging, particularly in settings where analytical resources and technical support are limited.

In response to these challenges, several digital platforms have been developed over the past decade, acting to facilitate outbreak detection and access to surveillance information. However, most systems focus on a specific part of the analytical process: collecting signals, displaying indicators, monitoring events, or providing predefined dashboards. Few of them support the full workflow that public health professionals routinely perform [5], from data discovery and exploration to analysis, modeling, visualization, and report generation.

Rather than focusing solely on outbreak detection, epidemic intelligence in practice involves a broader set of activities. Public health professionals must gather information from multiple sources, analyze trends, develop and evaluate models, interpret results, and communicate findings to support decision-making [6]. In this work, the term **Generalized Epidemic Intelligence (GEI)** is defined as the end-to-end analytical process spanning data discovery, analysis, modeling, visualization, and communication of findings to support public health decision-making.

Natural language interfaces have transformed how people interact with information. Today, users can ask complex questions without knowing how a database is structured, how a search engine ranks results, or how information is stored behind the scenes [7]. However, information retrieval is only the first step in the broader process of epidemiological analysis. An epidemiologist investigating an outbreak may need to compare trends across regions, explore changes over time, identify potential drivers of transmission, generate maps and visualizations, evaluate intervention scenarios, and communicate findings to different audiences. Even when the necessary data are publicly available, these tasks often require multiple software tools and a combination of technical skills that extend beyond epidemiology itself [8].

The challenge, therefore, is not simply providing access to information, but to support the analytical process that transforms information into evidence. This raises an important question: *Can natural-language systems do more than answer questions? Can they support the broader workflows involved in epidemiological analysis and public health decision-making?* [7] To explore this question, we developed EpidBot, a platform designed to support epidemiological analysis through natural language interaction.

Rather than focusing on a single task, such as information retrieval or outbreak detection, EpidBot was designed to orchestrate the broader analytical workflows routinely performed in public health practice.

The platform brings together multiple public health data sources within a single environment and combines them with tools for data exploration, visualization, statistical analysis, mathematical modeling, and report generation. Users can traverse the entire analytical workflow — from formulating a question to obtaining evidence-based results — within a single conversational interface, and it has been piloted in collaboration with governmental and international public health organizations.

In this article, we describe EpidBot and illustrate its use through representative analytical tasks. These include surveillance data exploration, multi-source epidemiological analyses, automated report generation, and the construction of a mathematical model for Ebola virus disease in the Democratic Republic of the Congo. Through these examples, we examine how natural language interfaces can support epidemiological work while maintaining transparency, reproducibility, and clear data provenance.

### Objectives

This article presents the conceptual framework, architecture, and implementation of EpidBot, a platform designed to support Generalized Epidemic Intelligence(GEI) through natural language interaction. We describe the GEI framework, the system architecture, the data integration strategy, and illustrate the platform through representative epidemiological investigations spanning surveillance, data integration, epidemiological analysis, and modeling.

## Materials and methods

### Generalized Epidemic Intelligence

Traditional epidemic intelligence systems have primarily focused on surveillance activities, including data collection, event detection, and situational awareness [9] [10] [11]. However, modern public health decision-making often requires a broader set of analytical activities that extend beyond surveillance alone, including evidence synthesis, statistical analysis, mathematical modeling, scenario evaluation, and technical report generation [12].

To address this gap, we propose the concept of *Generalized Epidemic Intelligence* (GEI), defined as an integrated analytical framework that transforms epidemiological questions into evidence-informed decision-support products through a sequence of interconnected analytical processes. Rather than treating surveillance, analysis, modeling, and reporting as independent tasks, GEI views them as components of a unified workflow.

The novelty of GEI lies in bringing together activities that are often performed independently in routine public health practice. Instead of treating surveillance, analysis, modeling, and reporting as separate tasks, GEI organizes them into a single analytical workflow that starts with a natural-language question and ends with a decision-support product.

As illustrated in Figure 1, the process begins with a public health question expressed in natural language. Relevant epidemiological data and scientific evidence are then retrieved and synthesized, forming the basis for exploratory analyses and mathematical modeling. The resulting analyses are communicated through technical reports, visualizations, projections, and other products intended to support public health decision-making.

**Fig 1.**
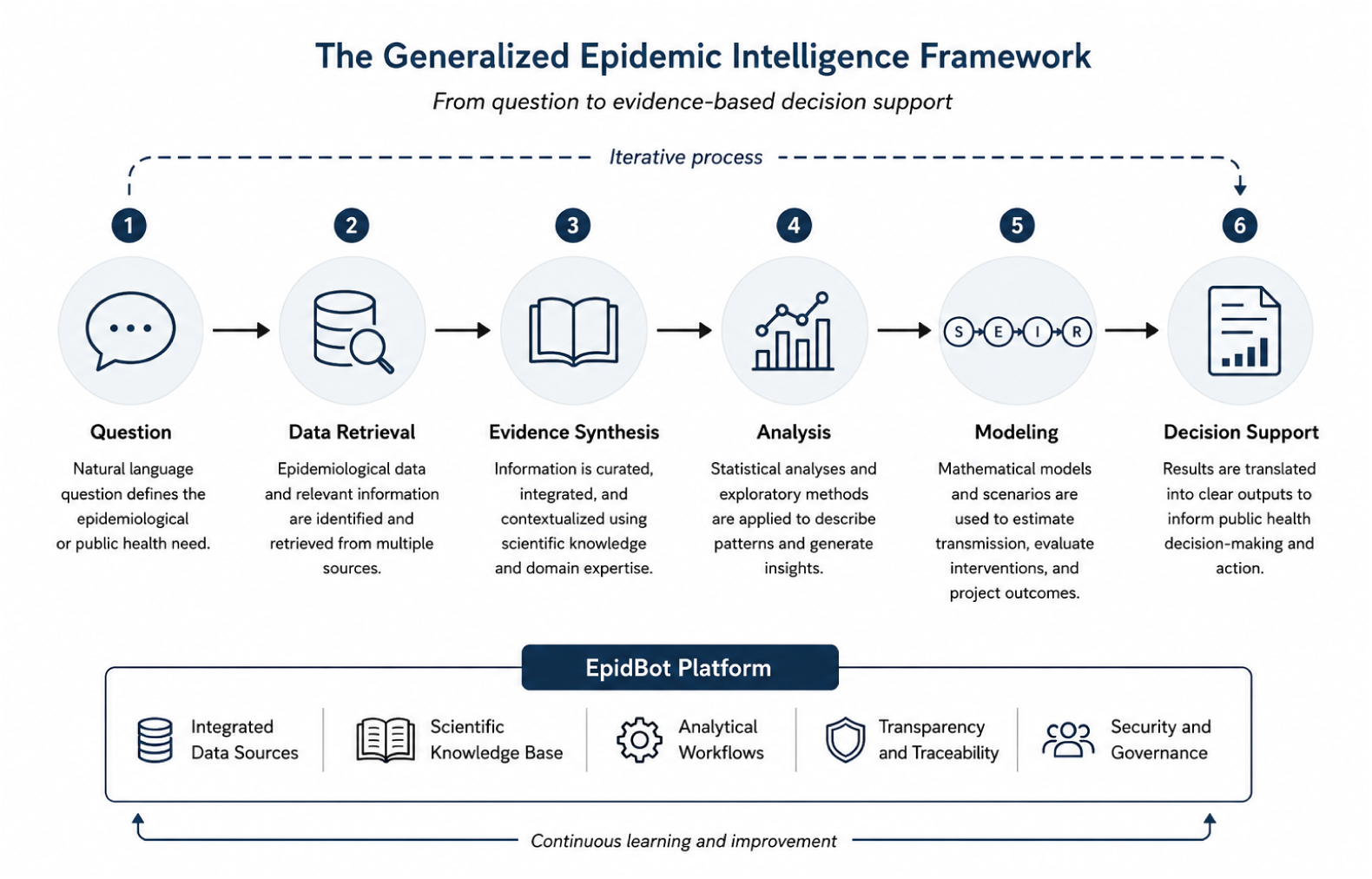
Conceptual framework for Generalized Epidemic Intelligence (GEI). Public health questions initiate an analytical workflow that integrates data retrieval, evidence synthesis, epidemiological analysis, mathematical modeling, and decision-support generation. The framework emphasizes the transformation of heterogeneous data and scientific knowledge into actionable public health intelligence through a unified analytical process.

The GEI framework is organized around five foundational components: (i) integrated data sources, which provide access to heterogeneous epidemiological information; (ii) a scientific knowledge base, which supplies contextual evidence from the scientific literature and public health guidance from reference documents from governments and renowned public health institutions such as the CDC, ECDC, Fiocruz, and others; (iii) analytical workflows, which transform data into epidemiological insights; (iv) transparency and traceability mechanisms, which expose analytical steps, data provenance, and scientific references; and (v) security and governance procedures, which ensure responsible use of data and analytical outputs.

Within this framework, EpidBot serves as an operational implementation of GEI, providing a analytical environment through which epidemiological workflows can be executed, documented, and reproduced. The following section describes the system architecture that supports this implementation.

### System architecture

EpidBot is a conversational platform for epidemiological analysis that combines natural language interaction with automated data retrieval, analytical workflows, and report generation. Natural language requests are interpreted by a large language model that coordinates access to external data sources, scientific knowledge repositories, and computational tools.

For each user request, the system decomposes the task into a sequence of analytical operations, including data retrieval, transformation, statistical analysis, visualization, mathematical modeling, and report generation. These operations are executed through a collection of specialized tools that can be dynamically combined according to the requirements of each epidemiological question [7, 13].

While the GEI framework defines the conceptual process through which epidemiological questions are transformed into decision-support products, EpidBot provides the computational infrastructure required to operationalize this process. As illustrated in Figure 2, the platform integrates heterogeneous data sources, scientific knowledge repositories, and analytical services within a unified analytical environment.

**Fig 2.**
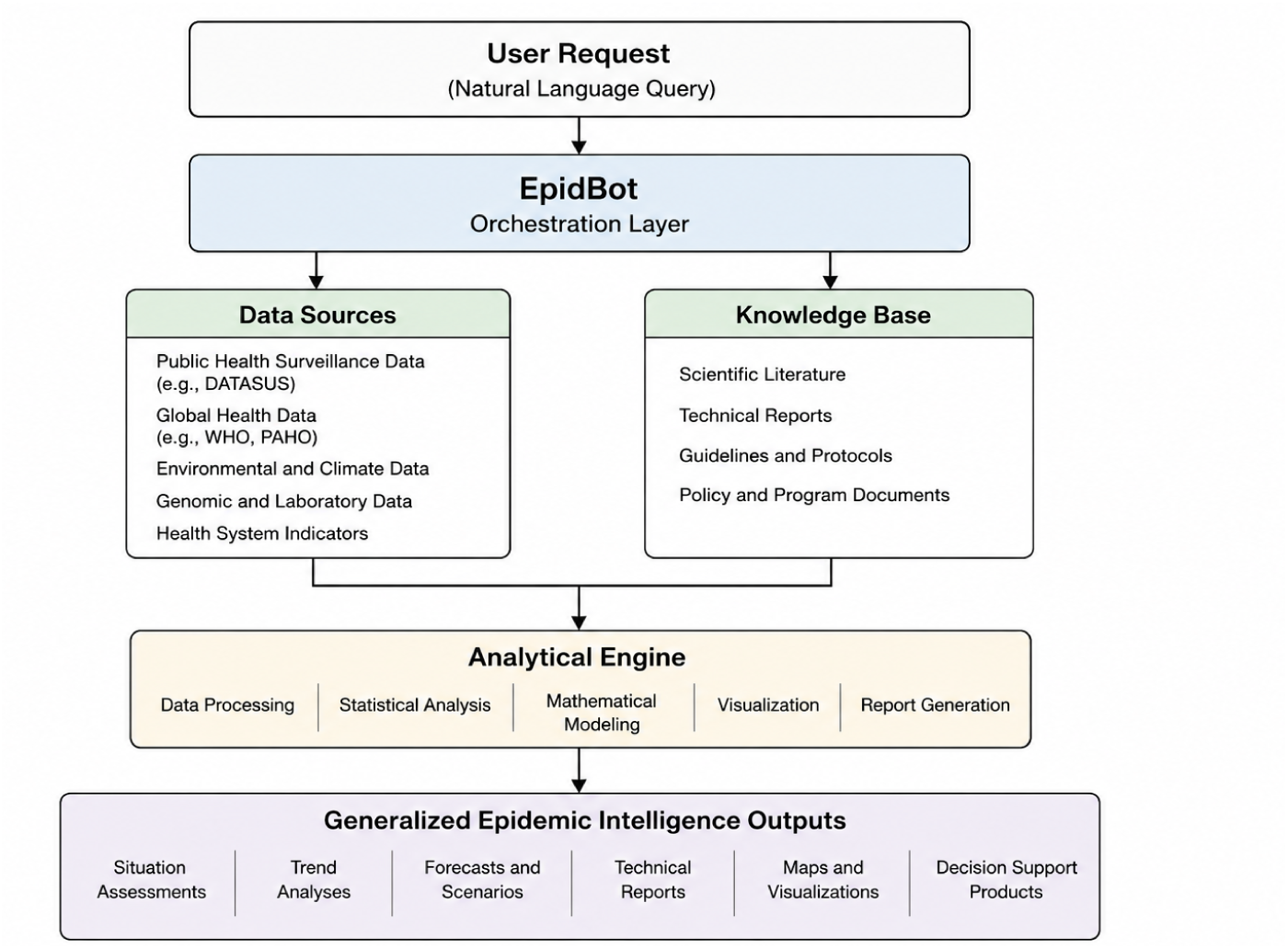
Overview of the EpidBot architecture. The platform integrates epidemiological data sources, scientific knowledge repositories, and computational tools within a unified analytical environment. User requests are transformed into analytical processes that support data exploration, statistical analysis, mathematical modeling, visualization, and report generation, producing outputs intended to support epidemiological assessment and public health decision-making.

The architecture shown in Figure 2 is organized into four functional layers that collectively support data integration, analytical execution, visualization, and knowledge management:

#### Layer 1: Epidemiological Data Integration

A central challenge in epidemiological analysis is that relevant information is rarely located in a single repository. Public health questions often require the integration of surveillance records, demographic indicators, environmental data, genomic information, health service statistics, and scientific evidence originating from multiple institutions and geographic scales [14, 15].

To address this challenge, the first architectural layer of EpidBot provides access to a diverse ecosystem of epidemiological data sources spanning local, national, regional, and global levels. The platform integrates information from more than a dozen public data providers, including Brazilian health information systems maintained by the Ministry of Health, regional repositories maintained by the Pan American Health Organization (PAHO), and global resources provided by organizations such as the World Health Organization (WHO), the World Bank, and the European Centre for Disease Prevention and Control (ECDC). Additional sources include climate and environmental datasets, genomic surveillance repositories, and disease-specific monitoring platforms [16–18].

Examples of integrated data sources spanning local, national, regional, and global levels are summarized in Table 1.

**Table 1.** Representative examples of integrated data sources integrated into EpidBot across different geographic scales. They are illustrative and do not represent the complete inventory of integrated sources.

| Level | Examples of Sources and Data |
| --- | --- |
| Local | Mosqlimate, Infodengue, and GADM, providing disease incidence, epidemic alerts, climate indicators, mosquito surveillance, and administrative boundaries. |
| National | DATASUS systems (SINAN, SIM, SINASC, SIH, SIA, PNI, and CNES), providing notifiable diseases, mortality, births, hospitalizations, outpatient procedures, immunization, and health infrastructure data [19]. |
| Regional | PAHO repositories, including health indicators, immunization coverage, malaria surveillance, and regional public health statistics. |
| Global | WHO, World Bank, ECDC, Pathoplexus, and the Malaria Atlas Project, providing outbreak reports, health indicators, respiratory surveillance, genomic sequences, and environmental datasets. |

Rather than requiring users to locate, access, and harmonize these resources independently, EpidBot provides a unified access layer through which heterogeneous datasets can be retrieved and analyzed within a common analytical environment. This approach enables epidemiological investigations that combine information across domains and geographic scales while reducing the technical barriers traditionally associated with multi-source data integration.

The integration layer is extensible, allowing new data sources to be incorporated as analytical needs evolve and additional public health datasets become available.

#### Layer 2: Analytical Execution

Access to data alone is rarely sufficient for epidemiological decision-making. Public health professionals must still identify trends, compare populations, investigate potential drivers of disease, and, in some situations, estimate future scenarios. These tasks often require multiple analytical tools, programming skills, and considerable time to execute.

The second architectural layer is responsible for transforming retrieved data into epidemiological evidence. To support this process, EpidBot provides an analytical environment capable of exploratory data analysis, statistical evaluation, visualization, and mathematical modeling. Depending on the question being investigated, analyses may involve temporal trends, demographic stratification, spatial comparisons, seasonality assessments, climate–health relationships, outbreak characterization, or epidemiological projections.

A distinguishing characteristic of EpidBot is that analytical procedures are not restricted to predefined dashboard components. Instead, analytical workflows are assembled according to the epidemiological question being investigated. Different questions may therefore require different combinations of datasets, statistical methods, visualizations, and reporting strategies.

These capabilities are supported by an integrated computational environment that combines database querying, statistical computing, and scientific programming tools, enabling both routine epidemiological analyses and more specialized investigations.

#### Layer 3: Visualization and Spatial Intelligence

Epidemiological analyses are often difficult to interpret from tables alone.

Identifying geographic clusters, temporal trends, seasonal patterns, or differences between populations typically requires visual exploration of the data [20].

For this reason, visualization is integrated directly into the analytical workflow. Rather than producing numerical outputs alone, EpidBot generates figures that help contextualize epidemiological findings and communicate results to decision-makers. Depending on the analysis, outputs may include epidemic curves, temporal trend plots, comparative visualizations, distribution summaries, and geographic maps.

Spatial analysis is particularly important in public health because many epidemiological questions are inherently geographic. Disease burden, access to healthcare, environmental exposures, and outbreak dynamics often vary substantially across locations [21, 22]. EpidBot therefore incorporates geographic reference information that allows epidemiological indicators to be linked automatically to administrative boundaries and visualized at different spatial scales.

These visualizations are generated as part of the analytical process and incorporated directly into technical reports, allowing data, analyses, and interpretations to be presented within a single decision-support product [23, 24].

#### Layer 4: Knowledge Integration and Reproducibility

Epidemiological analyses frequently require information that extends beyond surveillance data alone. Interpretation of findings often depends on scientific literature, methodological guidance, disease-specific recommendations, and contextual public health knowledge.

To support this process, EpidBot incorporates a curated knowledge layer containing epidemiological reference materials, public health guidance documents, and technical documentation associated with integrated data sources. These resources can be consulted during an investigation to provide contextual information, support methodological decisions, and assist in the interpretation of analytical results.

The platform also includes a library of reusable analytical workflows derived from recurring epidemiological tasks. These workflows encode commonly used analytical procedures and reporting structures, facilitating consistency across investigations while preserving the flexibility required for different public health questions.

A central design principle of EpidBot is transparency and reproducibility. Users can inspect the data sources consulted, analytical procedures performed, code executed, and visualizations generated during an investigation [25, 26]. This allows analytical outputs to be independently verified and reproduced when necessary. When required information is unavailable, data quality is insufficient, or uncertainty cannot be resolved, the system explicitly reports these limitations rather than presenting unsupported conclusions.

Taken together, these components provide a unified framework through which epidemiological questions can be translated into analytical products that support public health decision-making.

In this sense, EpidBot serves as an operational implementation of the Generalized Epidemic Intelligence framework described in Figure 1.

### Data Integration Pipeline

Epidemiological investigations frequently require information from multiple sources that differ in geographic scale, format, update frequency, and accessibility [27, 28]. The datasets required for an investigation depend strongly on the question being addressed. For example, local health planning may require mortality records, demographic indicators, and health-service data, whereas outbreak investigations may require surveillance reports, mobility information, environmental data, and scientific literature.

Because epidemiological analyses are inherently exploratory, they are difficult to support through a fixed, preloaded database. Different investigations often require different combinations of datasets, analytical procedures, and geographic scales [29].

To address this challenge, EpidBot adopts a demand-driven data integration strategy. Rather than maintaining a centralized repository containing all available datasets, the platform identifies, retrieves, and harmonizes only the information required for a specific investigation [30]. Retrieved datasets are cataloged together with metadata describing their provenance, geographic coverage, temporal scope, and update characteristics, supporting both reproducibility and dataset reuse across future analyses [3, 31].

By integrating data acquisition, harmonization, and provenance tracking within a single workflow, the platform allows users to focus on epidemiological questions rather than the technical challenges of locating, accessing, and preparing data from multiple repositories. Additional details regarding the data integration infrastructure are provided in the Supplementary Materials, including representative examples of municipal health planning and climate-health investigations that illustrate the demand-driven integration strategy adopted by the platform (Supplementary Figures S1 and S2).An illustrative conversational workflow is provided in the Supplementary Materials.

### Natural-Language Epidemiological Analysis

Epidemiological investigations rarely follow a predefined sequence of analytical steps. Instead, analyses often evolve as new evidence emerges, requiring investigators to explore alternative hypotheses, examine different population groups, and integrate additional sources of information [32].

To support this process, EpidBot provides a conversational interface through which epidemiological questions can be expressed directly in natural language. Users are not required to write code, formulate database queries, or navigate multiple data repositories [33, 34]. Instead, data retrieval, analysis, visualization, and report generation are integrated within a single environment.An illustrative conversational workflow is provided in the Supplementary Materials.

A key feature of the platform is its ability to maintain context across multiple interactions. Rather than treating each request as an isolated query, EpidBot supports iterative investigations in which each analytical step builds upon previous results. This interaction pattern closely mirrors real-world epidemiological practice, where findings from one stage of an investigation often motivate additional analyses and refinement of hypotheses [34].

By integrating data access, analytical workflows, visualization, and reporting within a unified environment, the platform reduces many of the technical barriers traditionally associated with epidemiological analysis [35, 36]. Additional details regarding conversational workflow execution, task decomposition, ambiguity handling, multilingual interaction, and report generation are provided in the Supplementary Materials [33].

### Deployment Context

During the study period, EpidBot was made available to public health professionals, researchers, and technical staff operating across multiple domains of epidemiology and public health practice.

Users included epidemiologists, surveillance officers, public health managers, researchers, data scientists, biostatisticians, environmental health professionals, veterinary epidemiologists, physicians, nurses, endemic disease control agents, and professionals involved in genomic surveillance, vaccination research, health information systems, and climate-sensitive health programs [3]. The platform was also explored by technical personnel affiliated with municipal health services, academic institutions, the Brazilian Ministry of Health, and the World Health Organization.

The diversity of user profiles exposed the platform to a broad range of analytical needs, spanning local disease surveillance, outbreak investigation, health planning, epidemiological research, climate-health analysis, and epidemiological intelligence activities [28]. This diversity informed the development of the platform and contributed to the identification of recurring analytical workflows described in the Results section.

## Results

### From Question to Epidemiological Investigation

One of the first questions explored using EpidBot was deceptively simple: how did dengue behave in Rio de Janeiro and São Paulo during Brazil’s historic 2024 epidemic?

The question was submitted in natural language, without specifying which datasets should be consulted, which indicators should be compared, or which analytical methods should be applied. Answering it, however, required the integration of information from multiple sources and the synthesis of evidence spanning surveillance data, epidemiological reports, and scientific literature.

After receiving the request, EpidBot began by identifying potentially relevant data sources and retrieving available dengue surveillance information. The platform subsequently searched the scientific literature, consulted regional surveillance reports, and assembled the information required to construct a comparative assessment of the epidemic in both states (Figure 3).

**Fig 3.**
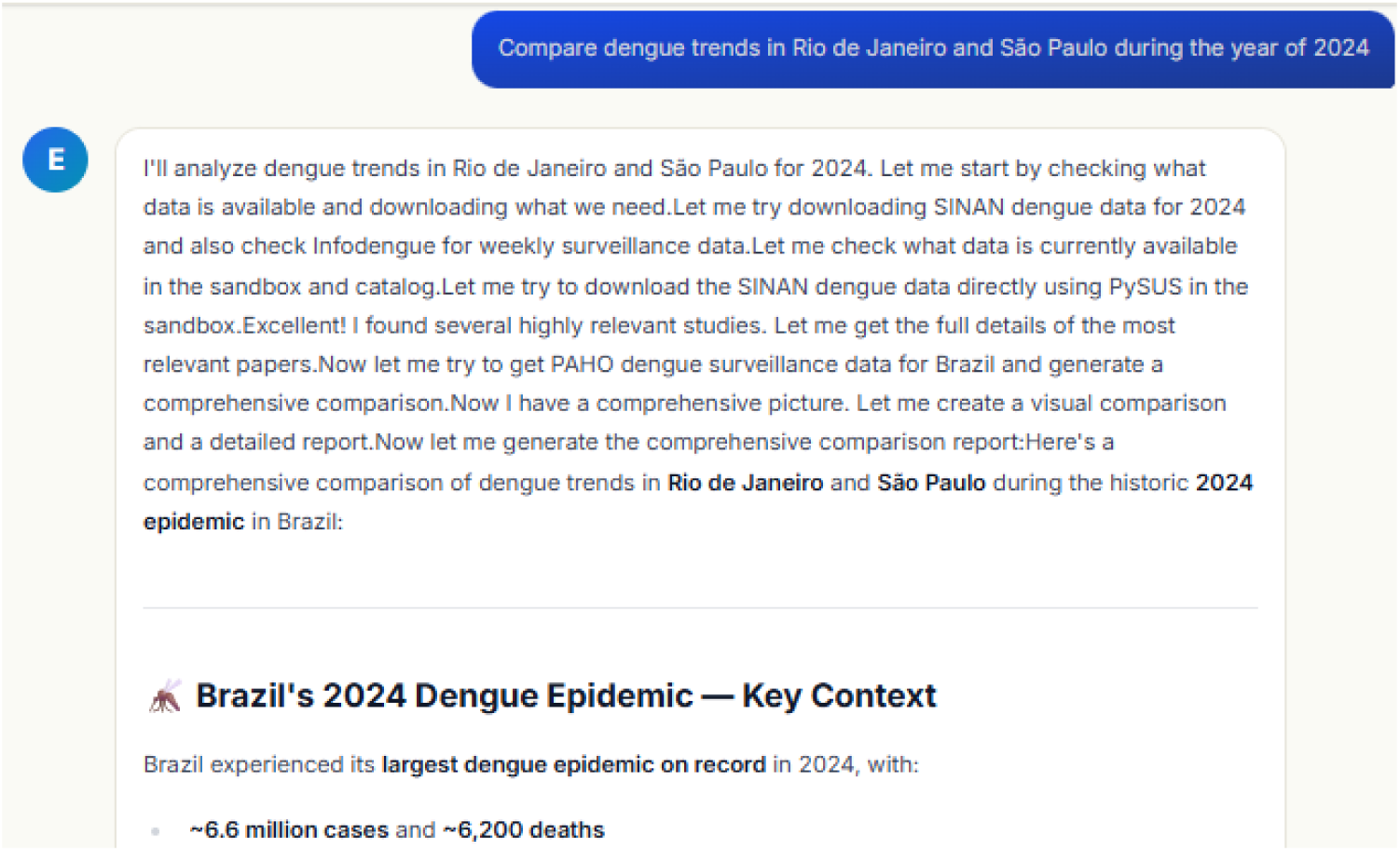
Example of a natural-language epidemiological question submitted to EpidBot. To answer the request, the platform identified relevant data sources, retrieved surveillance information, searched the scientific literature, and assembled the evidence required to support the analysis.

The resulting investigation highlighted important differences between the two states. São Paulo emerged as the national epicenter of the epidemic [37], experiencing an unprecedented disease burden and widespread circulation of DENV-1 and DENV-2 [38]. In contrast, although Rio de Janeiro was also heavily affected, the analysis identified evidence suggesting a substantial impact of Wolbachia-based vector control interventions, particularly in Niterói [39].

Rather than presenting these findings as isolated facts, the platform synthesized the retrieved evidence into an integrated analytical product that combined epidemiological indicators, contextual information, and findings reported in the scientific literature. Figure 4 shows an example of a summary generated during this process.

**Fig 4.**
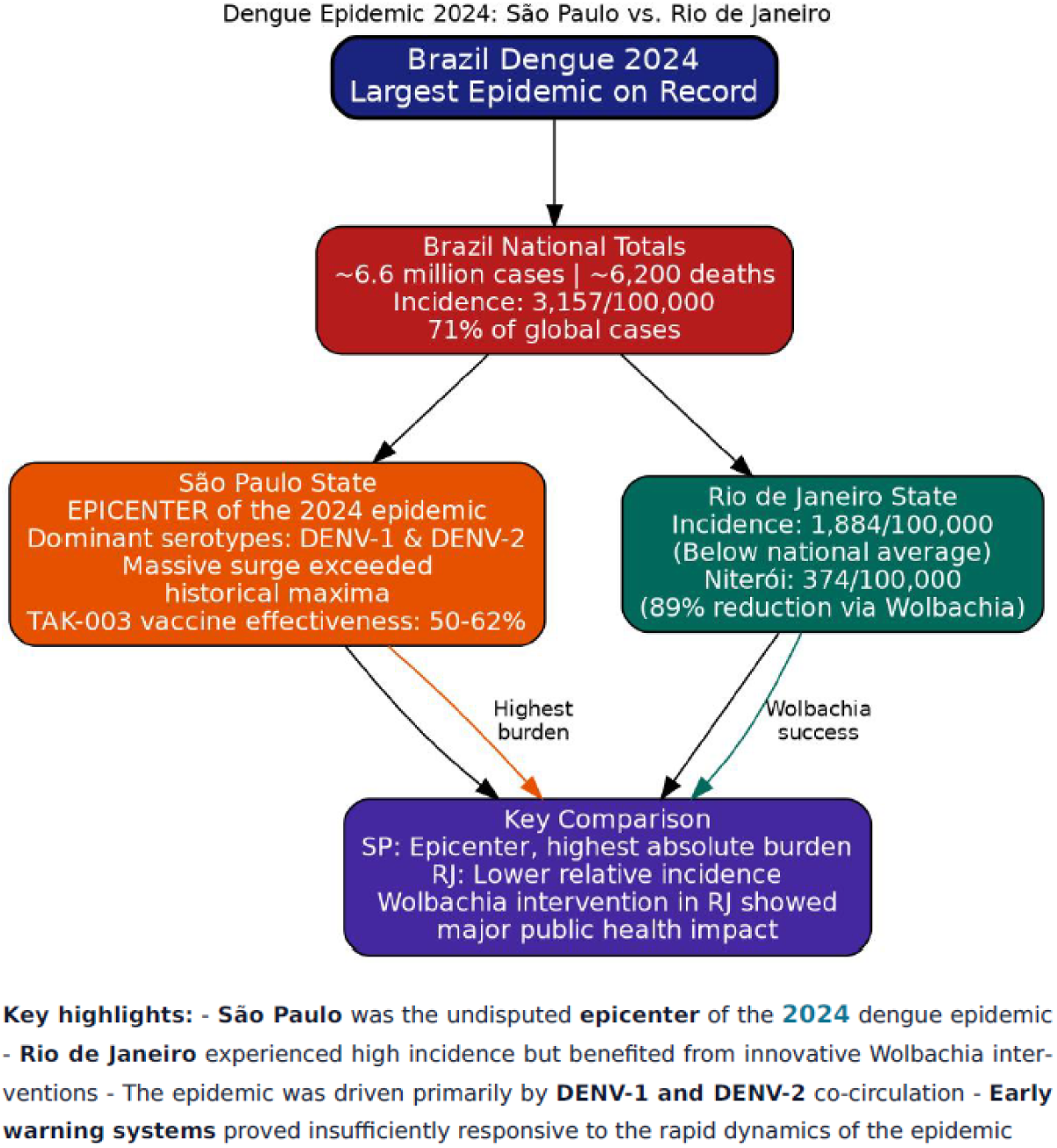
Example of a synthesized analytical output generated during the investigation. The summary combines epidemiological indicators and evidence retrieved from the literature to support the interpretation of observed differences between Rio de Janeiro and São Paulo during the 2024 dengue epidemic.

The investigation ultimately produced a structured technical report containing epidemiological context, comparative analyses, references to supporting evidence, and key public health interpretations. A complete version of the automatically generated report has been deposited in Zenodo to support transparency and reproducibility [40].

Although the original request consisted of a single question, the resulting workflow involved data acquisition, evidence retrieval, synthesis, visualization, and report generation. This example illustrates how natural-language interaction can be used to transform broad epidemiological questions into complete analytical investigations.

### Natural-Language Generation of Mathematical Epidemic Models

In addition to supporting epidemiological data exploration and situational assessments, EpidBot was able to construct mathematical models directly from natural-language requests. To evaluate this capability, we asked the platform to develop a transmission model for Ebola virus disease using information retrieved from outbreak reports and the scientific literature, including WHO outbreak reports [41].

Starting from a single user request, the platform identified key epidemiological characteristics of Ebola virus disease, assembled supporting evidence, formulated a compartmental transmission model, documented model assumptions, and generated a structured technical report. The resulting report included model equations, parameter definitions, intervention scenarios, simulation outputs, and analytical interpretations [42].

The generated workflow automatically identified epidemiologically relevant transmission pathways, including community, healthcare-associated, and funeral-related transmission routes [43] and translated them into a mathematical modeling framework. EpidBot subsequently defined a set of intervention scenarios representing alternative public health response strategies, including safe burial campaigns, ring vaccination, improvements in hospital care, and combinations of interventions (Table 2).

**Table 2.** Intervention scenarios automatically generated and evaluated during the Ebola modeling workflow.

| Scenario | Description | Modified Parameters |
| --- | --- | --- |
| S1: Baseline | No intervention (status quo) | All baseline values |
| S2: Safe Burial | Community engagement for SDB | $\theta$: 0.60 → 0.20 |
| S3: Ring Vaccination | rVSV-ZEBOV ring vaccination | $f_s$: 0.55 → 0.25, $\beta_c$: 0.30 → 0.10 |
| S4: Hospital Care | Improved case management | $f_h$: 0.30 → 0.70, $\gamma_h$: 1/7→1/5, $\mu_{dh}$: 0.03→0.01 |
| S5: Combined | All interventions simultaneously | All of the above |

After defining the intervention scenarios, the platform executed simulations and compared their projected epidemiological outcomes. Table 3 summarizes representative simulation results generated during the investigation.

**Table 3.** Simulation outcomes obtained under alternative intervention scenarios generated by EpidBot.

Simulation Results (365 days, N=100,000, I<sub>0</sub>=5)
| Metric | S1:Baseline | S2:SafeBurial | S3:Vaccination | S4:HospCare | S5:Combined |
| --- | --- | --- | --- | --- | --- |
| R<sub>0</sub> | 1.83 | 1.39 | 0.64 | 1.59 | 0.45 |
| Peak Active Infections | 2,317 | 827 | 5 | 1,403 | 5 |
| Peak Day | 222 | 365 | 10 | 266 | 8 |
| Total Cases | 73,621 | 22,869 | 13 | 60,504 | 8 |
| Total Deaths | 14,774 | 4,444 | 1 | 5,548 | 0 |
| CFR | 20.1% | 19.4% | 7.7% | 9.2% | 0.0% |

Substantial differences were observed across intervention strategies. Vaccination and combined intervention scenarios produced the largest reductions in projected transmission, cases, and mortality, whereas safe burial interventions alone also generated meaningful reductions compared with the baseline scenario [44]. These results illustrate how the platform can move beyond descriptive analyses to support the exploration of hypothetical intervention strategies and their potential epidemiological consequences.

This example demonstrates that natural-language interaction can support not only epidemiological data retrieval and evidence synthesis, but also the formulation of mathematical models, evaluation of alternative response strategies, and generation of structured analytical reports suitable for public health planning.

### Global Cholera Situation Assessment and Decision Support

Public health emergencies rarely arise from a single cause. Cholera outbreaks, for example, emerge from the interaction of environmental conditions, population displacement, water and sanitation infrastructure, health-system capacity, and broader social vulnerabilities [45] [46]. Understanding these interactions often requires navigating large volumes of information distributed across surveillance systems, humanitarian reports, climate datasets, and scientific literature.

To explore EpidBot’s ability to support this type of complex investigation, we conducted a global cholera assessment covering the period from 2023 to 2025. The objective was not limited to describing disease burden, but rather to understand where transmission was occurring, why certain regions were disproportionately affected, and which factors were most strongly associated with increased risk.

Starting from a natural-language request, the platform assembled information from multiple public sources and produced an integrated situation assessment. The resulting report combined surveillance indicators, country-level comparisons, temporal trends, environmental drivers, health-system characteristics, displacement patterns, and evidence from the scientific literature into a single analytical workflow.

Figure 5 presents an excerpt from the automatically generated report. Beyond summarizing global trends, the analysis highlighted the concentration of cholera burden in conflict-affected and resource-limited settings, while simultaneously identifying substantial variation in mortality patterns across countries. Rather than treating outbreaks as isolated events, the report contextualized them within broader regional and global dynamics [47].

**Fig 5.**
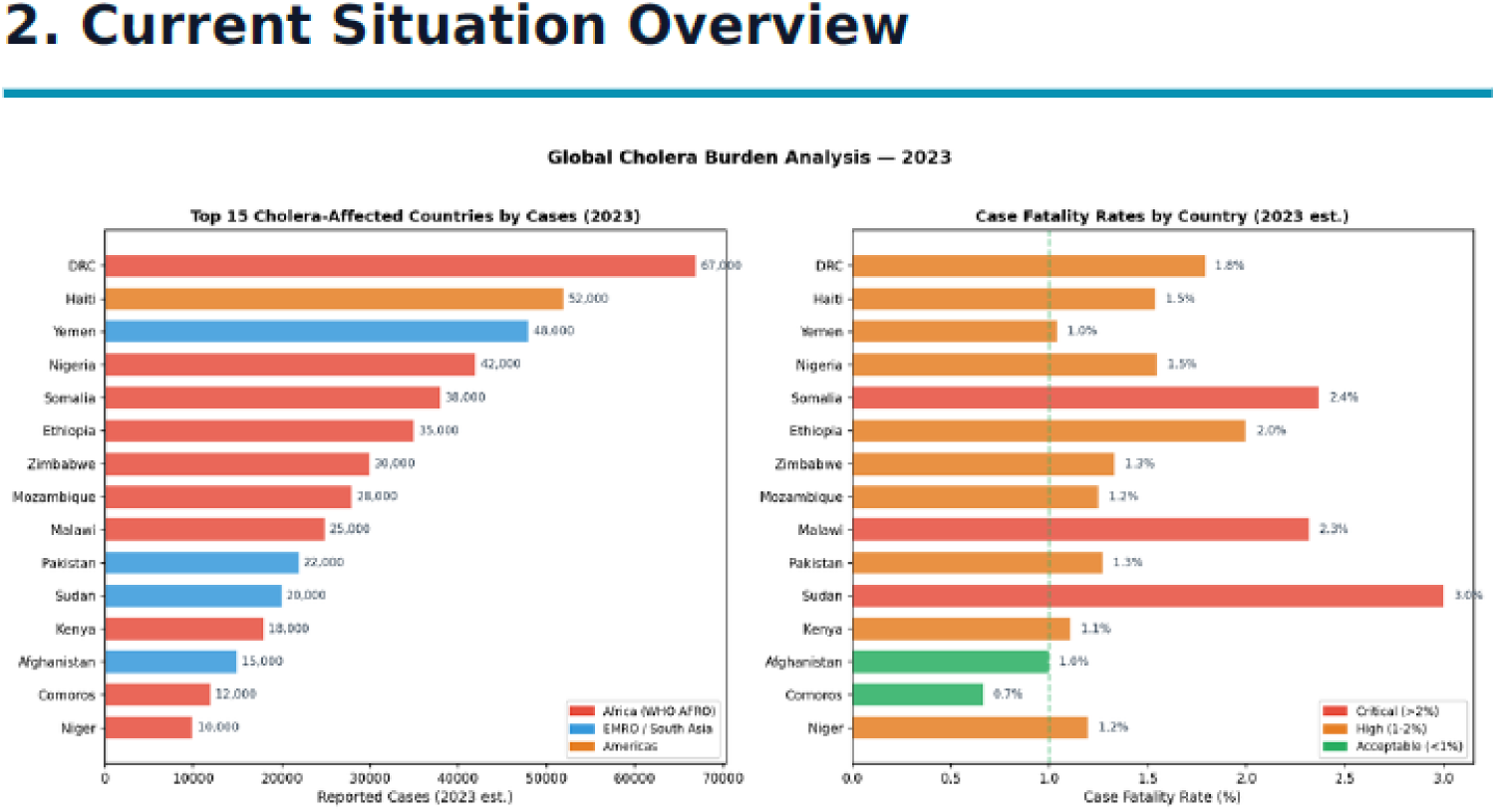
Overview of the global cholera assessment generated by EpidBot, including temporal trends and cross-country comparisons of disease burden and mortality.

A second layer of analysis focused on understanding the determinants of vulnerability. The platform automatically integrated indicators related to water and sanitation access, population displacement, health-system capacity, and broader socioeconomic conditions. Figure 6 illustrates how heterogeneous datasets can be combined to identify locations where outbreaks are more likely to emerge, persist, or overwhelm local response capacity.

**Fig 6.**
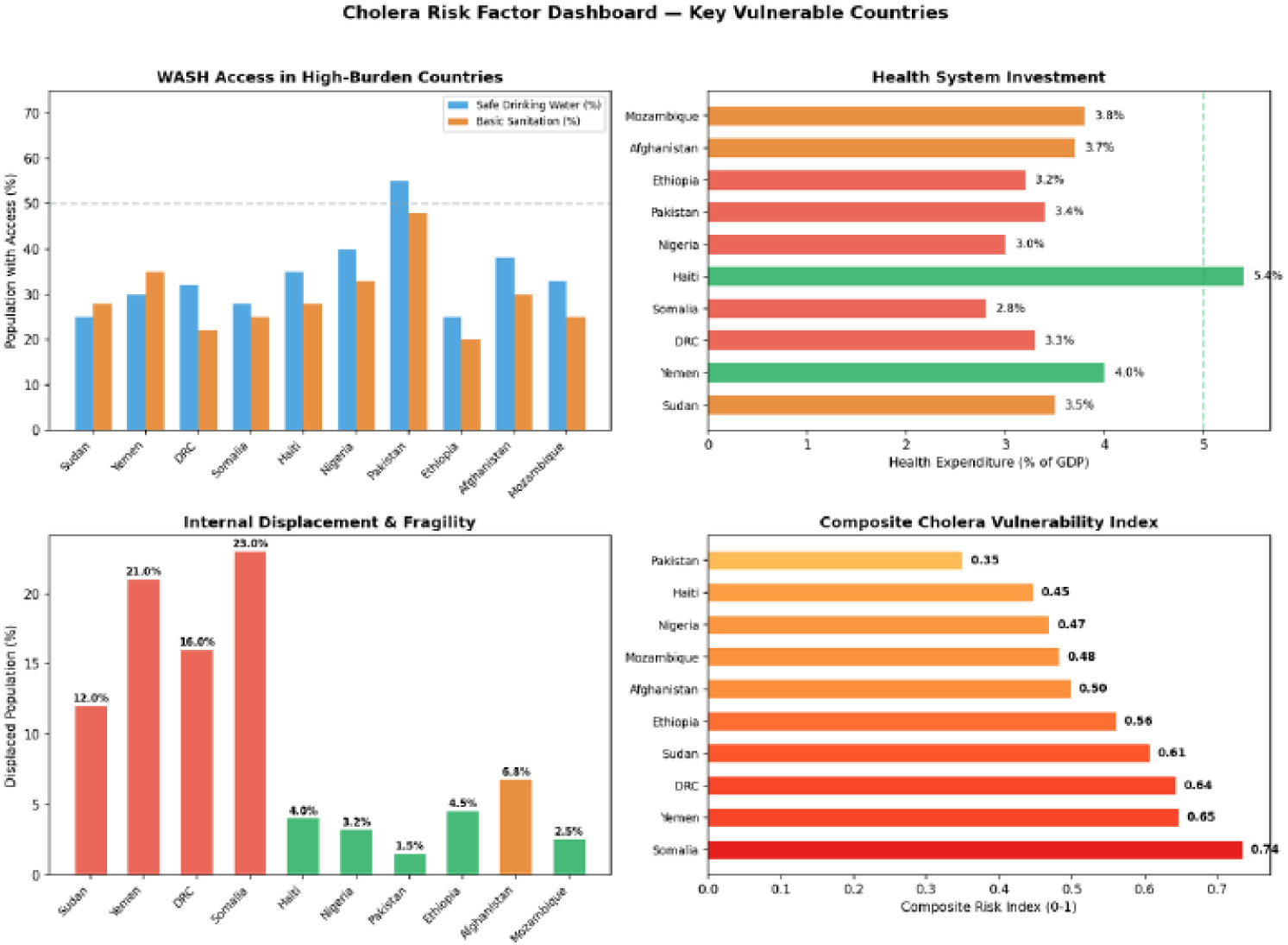
Multi-source vulnerability assessment generated during the cholera investigation, integrating epidemiological, environmental, infrastructural, and social indicators.

Importantly, the workflow did not end with descriptive analytics. After synthesizing the available evidence, EpidBot translated findings into operational recommendations, identifying priority regions, surveillance needs, and intervention opportunities. The platform also generated a conceptual framework describing how epidemiological surveillance, climate information, environmental monitoring, and artificial intelligence could be combined to support earlier outbreak detection and more effective response strategies (Figure 7).

**Fig 7.**
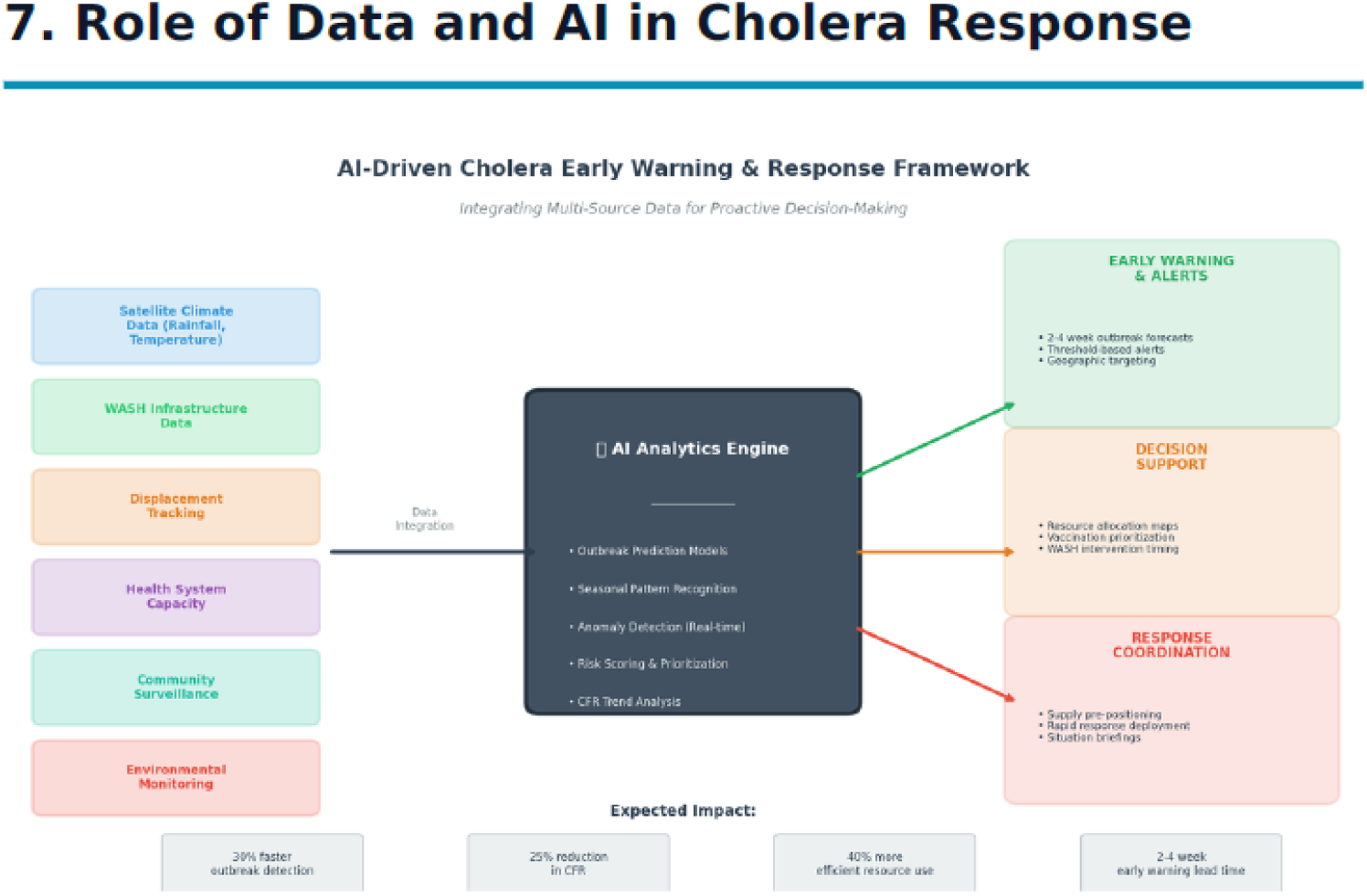
AI-enabled public health intelligence framework generated as part of the cholera assessment, illustrating how multiple information streams can support early warning and decision-making. cases, users interacted exclusively through natural language, while the platform dynamically assembled the analytical workflows required to answer each question.

This example illustrates a broader challenge facing public health systems. In many situations, the information required to support decision-making already exists, but is fragmented across institutions, formats, and disciplines. The bottleneck is often not the absence of data, but the difficulty of transforming dispersed information into coherent, actionable knowledge [29]. By allowing users to interact with diverse datasets, scientific evidence, and analytical tools through natural language, EpidBot seeks to reduce this gap and make advanced epidemiological intelligence more accessible to a wider range of public health professionals.

Taken together, these examples illustrate three distinct modes of operation supported by EpidBot. The dengue investigation demonstrates multi-source epidemiological analysis and evidence synthesis; the Ebola case illustrates natural-language-driven mathematical modeling and scenario evaluation; and the cholera assessment highlights the integration of heterogeneous data streams into decision-support products. Across all

## Discussion

This work suggests that natural-language interaction can support a broader range of epidemiological activities than those traditionally associated with epidemic intelligence. Rather than focusing exclusively on surveillance and event detection, EpidBot illustrates how epidemiological questions can be translated into evidence synthesis, exploratory analyses, mathematical modeling, and decision-support products within a unified analytical workflow.

The main contribution of this work is the introduction of the Generalized Epidemic Intelligence (GEI) framework. GEI does not introduce new epidemiological methods. Instead, it integrates activities that are often performed independently in routine public health practice. By connecting surveillance, evidence synthesis, data analysis, mathematical modeling, and report generation, GEI enables public health questions to be transformed into structured analytical investigations.

The examples presented in this work illustrate different ways in which this framework can be applied. The dengue investigation demonstrated the integration of multiple data sources and scientific evidence to support comparative epidemiological analyses. The Ebola example showed that natural-language requests can be translated into mathematical models and intervention scenarios. The cholera assessment highlighted how heterogeneous information streams can be combined to produce decision-support products that extend beyond disease surveillance alone. Although these examples address distinct public health problems, they all follow the same underlying principle: transforming a question into an analytical investigation.

The examples also highlight a broader challenge faced by public health systems. In many situations, the information required to support decision-making already exists, but remains fragmented across institutions, formats, and disciplines. The bottleneck is often not the absence of data, but the difficulty of transforming dispersed information into coherent and actionable knowledge. By allowing users to interact with heterogeneous datasets, scientific evidence, and analytical tools through natural language, EpidBot seeks to reduce this gap and expand access to epidemiological intelligence.

Importantly, EpidBot is not intended to replace epidemiological expertise. Public health investigations require contextual interpretation, critical assessment of evidence, and domain knowledge that cannot be delegated to automated systems. Rather, the platform should be viewed as an analytical support tool whose purpose is to augment the work of public health professionals and reduce some of the technical barriers traditionally associated with epidemiological analysis.

Future work will focus on expanding the range of supported epidemiological applications, strengthening integration with public health information systems, and evaluating the use of the platform across different operational contexts. Particular attention will be given to understanding how natural-language-driven analytical workflows can be safely incorporated into routine public health practice while maintaining transparency, traceability, and human oversight.

## Limitations

Several limitations should be considered when interpreting the results presented in this work.

First, EpidBot inherits some of the limitations associated with large language models. Although the platform relies on external tools and retrieved data rather than parametric memory, errors may still occur during the interpretation and synthesis of analytical results. For this reason, outputs intended to support operational public health decisions should always be independently verified by domain experts.

Second, the quality of the analyses depends directly on the availability, completeness, and timeliness of the underlying data sources. Surveillance systems are subject to reporting delays, underreporting, changes in case definitions, and inconsistencies that vary across countries and over time. EpidBot can identify some of these limitations, but it cannot fully correct them.

Third, EpidBot is designed as an analytical support tool and not as a substitute for epidemiological expertise. Statistical associations should not be interpreted as causal relationships without appropriate epidemiological assessment, and findings generated by the system should always be interpreted within their public health context.

Fourth, full reproducibility cannot always be guaranteed. Although data retrieval, code execution, and analytical outputs can be documented and exported, the sequence of analytical steps selected during an investigation may vary slightly across sessions due to the stochastic nature of large language models.

Finally, the platform has been most extensively tested using Brazilian epidemiological datasets. Although international data sources are supported, differences in data dictionaries, reporting standards, and administrative boundaries may affect analytical consistency across countries.

## Conclusion

This work introduced the concept of Generalized Epidemic Intelligence (GEI) and presented EpidBot as its operational implementation. We showed how natural-language interaction can be used to transform epidemiological questions into structured analytical investigations that integrate data retrieval, evidence synthesis, statistical analysis, mathematical modeling, visualization, and report generation.

The examples presented in this work demonstrate that epidemic intelligence can be extended beyond traditional surveillance activities. Rather than functioning solely as a mechanism for event detection, GEI provides a framework through which epidemiological questions can be translated into decision-support products that integrate multiple sources of evidence.

EpidBot is not intended to replace epidemiological expertise. Instead, its purpose is to reduce technical barriers, facilitate access to heterogeneous sources of information, and augment the analytical capacity of public health professionals.

Future work will focus on expanding the range of supported epidemiological applications, strengthening integration with public health information systems, and evaluating the use of the platform across different operational contexts. Together, GEI and EpidBot suggest a new paradigm in which natural-language interaction becomes an interface for epidemiological investigation, enabling broader access to analytical capabilities that have traditionally required specialized technical expertise.

## Supporting information

## Data Availability

Data used in this study were obtained from publicly available sources referenced throughout the manuscript. Representative EpidBot-generated analytical reports have been deposited in Zenodo and are publicly accessible through the links provided below.

https://zenodo.org/doi/10.5281/zenodo.20706800

https://zenodo.org/records/20634292

https://zenodo.org/doi/10.5281/zenodo.20706786

## Acknowledgments

We thank the WHO Regional Office for the Eastern Mediterranean (WHO EMRO) for facilitating the pilot deployment of EpidBot and for the valuable contributions of its public health professionals throughout the evaluation process.

We also thank the Municipality of Joinville, Brazil, for its collaboration and for providing a real-world environment for testing and refining the platform.

Finally, we are grateful to the more than 40 researchers, epidemiologists, surveillance officers, public health professionals, and health managers who participated in the pilot activities and provided continuous feedback that helped shape the development and evaluation of EpidBot.

## Competing interests

The authors have read the journal’s policy and have the following competing interests: Flavio Codeço Coelho (FCC) and Joyce Figueiró Braga (JFB) are co-founders of Kwar-AI, a company focused on artificial intelligence solutions for public health that commercially develops EpidBot. This does not alter our adherence to PLOS policies on sharing data and materials.

## Author contributions

**Conceptualization:** FCC, JFB.

**Methodology:** FCC, JFB.

**Software:** FCC.

**Investigation:** FCC, JFB.

**Validation:** FCC, JFB.

**Writing – original draft:** JFB, FCC.

**Writing – review & editing:** FCC, JFB, BL.

**Supervision:** FCC.

**Project administration:** FCC, JFB.

## Notes

### Competing Interest Statement

Flavio Codeco Coelho (FCC) and Joyce Figueiro Braga (JFB) are co-founders of Kwar-AI, a company that develops EpidBot, the platform described in this manuscript. This does not alter the authors' commitment to transparency, data availability, and scientific integrity.

### Author Declarations

This study used only publicly available data and scientific literature that were accessible prior to the initiation of the study. Epidemiological analyses were based on publicly available surveillance data, reports, and publications obtained from official public health institutions and scientific repositories, including DATASUS (https://datasus.saude.gov.br), InfoDengue (https://info.dengue.mat.br), the World Health Organization (https://www.who.int), the Global Task Force on Cholera Control (https://www.gtfcc.org), and peer-reviewed scientific literature indexed in PubMed (https://pubmed.ncbi.nlm.nih.gov). No individual-level identifiable data, restricted-access datasets, or datasets requiring institutional approval, registration, or application procedures were used.

